# Real-time monitoring of COVID-19 dynamics using automated trend fitting and anomaly detection

**DOI:** 10.1101/2020.09.02.20186502

**Authors:** Thibaut Jombart, Stéphane Ghozzi, Dirk Schumacher, Quentin J Leclerc, Mark Jit, Stefan Flasche, Felix Greaves, Tom Ward, Rosalind M Eggo, Emily Nightingale, Sophie Meakin, Oliver J Brady, Centre for Mathematical Modelling of Infectious Diseases COVID-19 Working Group, Graham F Medley, Michael Höhle, W John Edmunds

**Author notes:** **Corresponding authors: Thibaut Jombart** Department of Infectious Disease Epidemiology London School of Hygiene & Tropical Medicine Keppel Street London WC1E 7HT United Kingdom **Stéphane Ghozzi** Department of Epidemiology Helmholtz Centre for Infection Research Inhoffenstraße 7 38124 Braunschweig/Brunswick Germany. Contributed equally to the manuscript (Thibaut Jombart, Stéphane Ghozzi). CMMID COVID-19 Working Group gave input on the method, contributed data and provided elements of discussion. The following authors were part of the Centre for Mathematical Modelling of Infectious Disease 2019-nCoV working group. Each contributed in processing, cleaning and interpretation of data, interpreted findings, contributed to the manuscript, and approved the work for publication.

## Abstract

As several countries gradually release social distancing measures, rapid detection of new localised COVID-19 hotspots and subsequent intervention will be key to avoiding large-scale resurgence of transmission. We introduce ASMODEE (Automatic Selection of Models and Outlier Detection for Epidemics), a new tool for detecting sudden changes in COVID-19 incidence. Our approach relies on automatically selecting the best (fitting or predicting) model from a range of user-defined time series models, excluding the most recent data points, to characterise the main trend in an incidence. We then derive prediction intervals and classify data points outside this interval as outliers, which provides an objective criterion for identifying departures from previous trends. We also provide a method for selecting the optimal breakpoints, used to define how many recent data points are to be excluded from the trend fitting procedure. The analysis of simulated COVID-19 outbreaks suggest ASMODEE compares favourably with a state-of-art outbreak-detection algorithm while being simpler and more flexible. We illustrate our method using publicly available data of NHS Pathways reporting potential COVID-19 cases in England at a fine spatial scale, for which we provide a template automated analysis pipeline. ASMODEE is implemented in the free R package *trendbreaker*.

## INTRODUCTION

After a fast initial spread worldwide and large-scale epidemics in many affected countries, the trajectory of the COVID-19 pandemic is changing. In a number of severely affected countries, strong mitigation measures such as various forms of social distancing have slowed national epidemics and in many cases brought the epidemic close to control (1–3). However, in the absence of widespread, long-lasting immunity through vaccination or natural infection (4–6), these respites are most likely temporary, and further relapses, in the form of localised outbreaks or nation-wide resurgence, remains highly likely and a very serious threat.

In the UK, a “lockdown” was implemented on 23 March 2020, and gradually relaxed from the beginning of June 2020, at which point about 300,000 confirmed COVID-19 cases and 40,000 deaths had been reported (7). Unfortunately, the risk of local flare-ups was illustrated soon after, as increased case incidence in Leicester resulted in the city being put under lockdown again on 29 June 2020 (8). Similarly, increased restrictions were imposed in Blackburn on 9 August 2020.

In order to prevent large-scale relapses, localised COVID-19 hotspots *(i.e*. places with high levels of transmission) need to be detected as soon as cases occur and contained as early as possible. For such detection to be optimal, COVID-19 dynamics need to be monitored at a small spatial scale, requiring daily surveillance of multiple time series of case incidence, and prompt detection of ongoing increases. Disease surveillance algorithms have been designed for such purpose (9–12), although many of them are tailored to detecting either seasonal or point-source outbreaks and may be most effective when trained on years of weekly incidence data (e.g. Farrington algorithm (13,14)). However, careful implementation of such algorithms has proved useful as a backbone for setting up automated disease surveillance systems for endemic diseases (15).

Here, we introduce ASMODEE (Automated Selection of Models and Outlier DEetection for Epidemics), an algorithm for detecting ongoing changes in COVID-19 incidence patterns. In order to characterise potentially very different dynamics in case incidence across a large number of locations, our approach implements a flexible time series framework using a variety of models including linear regression, generalised linear models (GLMs) or Bayesian regression. ASMODEE first identifies past temporal trends using automated model selection, and then uses outlier detection inspired by classical Shewhart control-charts to signal recent anomalous data points. We used this approach to design automated surveillance pipelines which monitor changes in potential COVID-19 cases reported through an online and telephone hotline used in England, the NHS Pathways system, which includes calls made to 111/999 as well as reports made through the 111-online system. We conducted the analysis at the level of Clinical Commissioning Groups (CCGs), small area divisions used for healthcare management in England’s NHS, with an average of 226,000 people each. One advantage of the NHS Pathways system is that reports occur with little delay, because no confirmatory diagnostic tests are involved; on the other hand it has the usual sensitivity and specificity issues of a syndromic surveillance system.

We used simulations to evaluate the potential of ASMODEE for detecting changes in incidence patterns. COVID-19 incidence dynamics were simulated using a branching process model with realistic estimates of the time-varying reproduction number *(Rt)* and serial interval, under four scenarios: steady state *(Rt* close to 1), relapse, lockdown and flare-up following low levels of transmission. For comparison, we also applied the modified Farrington algorithm (14), a standard method designed for the detection of point-source outbreaks and used in many public health institutions (9). We computed a variety of scores such as probability of detection, sensitivity and specificity for two configurations of ASMODEE. In addition, we show that when applied to NHS 111/999 calls data, ASMODEE would have enabled the early detection of the flare-ups in Leicester and Blackburn with Darwen in June and July 2020, respectively. We propose that ASMODEE may be a useful, flexible complement to existing outbreak detection methods for designing disease surveillance pipelines, for COVID-19 and other diseases.

All source code necessary to run ASMODEE, analyse and visualise results, implement the NHS Pathways pipeline, and reproduce the analyses is open and freely available under MIT license (see ‘availability’ sections in ‘Methods’).

## MATERIAL AND METHODS

### Automated Selection of Models and Outlier DEtection for Epidemics (ASMODEE)

#### General algorithm

ASMODEE is designed for detecting recent departures/aberrations from past temporal trends in univariate time series. The response variable typically represents case counts, but it can readily accommodate other response variables such as incidence rates or case fatality ratios. We are interested in classifying outliers in the most recent time points of the available time series. Our approach can be broken down into the following steps:

1. Partition the analysed time series into two complementary time windows: a *calibration window*, excluding the most recent *k* data points; and a *prediction window*, made of the last *k* data points. The value of *k* is either fixed by the user, or its ‘optimal’ value can be determined using heuristics (see ‘*Setting the value of k’* below). The corresponding datasets are referred to as the *calibration*, and *prediction set*, respectively.
2. Fit a range of user-defined time series models to the calibration set, and retain the best-predicting or best-fitting model (see ‘*Selecting the best model*’ below); here we describe implementations with linear regressions, various GLMs (including Poisson and Negative Binomial models), and Bayesian regression models.
3. Derive prediction intervals at a given *alpha* threshold (defaulting to 5%) for all data points, included in the prediction set. This threshold is equal to the expected proportion of data points that will be classified as outliers if all points followed the same temporal trend.
4. Identify outliers as data points lying outside the prediction interval; the number of data points showing a marked *increase* from past trends in the prediction set can be used to design an alarm system. Similarly a *decrease* in trend can be seen in data points lying below the lower bound of the prediction interval. Points within the interval can be considered as *normal, i.e*. belonging to the recent trend.

#### Selecting the ‘best’ model

The literature on model selection offers a plethora of methods for selecting models fulfilling a given optimality criteria in ‘traditional’ statistics (16–19) as well as in machine learning (20–22). To reflect some of this diversity, we have implemented two different approaches for model selection in ASMODEE. Note that both approaches use only the calibration window, and ignore data from the prediction window.

The first approach is *K*-fold cross-validation, in which the data is randomly broken into *K* partitions of roughly equal sizes (the “folds”) (23). Each fold is in turn used for a round of cross-validation: the *K*-1 other folds are used as a *training* set, to fit the model, while the remaining fold is used as a *testing* set, to assess the quality of the model predictions. By default, we use *K* = *N*, where *N* is the number of data points in the calibration window. Discrepancies between model predictions and observed data of the testing set can be measured using various metrics. Here, we chose the Root Mean Square Error (RMSE), because of its wide use in statistical modelling and ease of computation. RMSE is summed over all rounds of cross-validation to characterise a given model. The model with the lowest total RMSE is retained as the ‘best’ model. Note that this metric gives equal weights to positive and negative residuals, and thus implicitly assumes that the magnitude of deviations from the model (under and over estimation) is similar. Other metrics could be used with low counts, where negative residuals are typically smaller (as case counts cannot be negative). When *K* < *N*, further accuracy of measurement can be gained by repeating the cross-validation procedure several times, each time using a different set of random folds, called repeated *K*-fold cross-validation.

This approach is likely to select models with good predictive ability (in the sense of minimum RMSE), but can be computationally intensive when evaluating multiple models over many datasets. As an alternative, we also implemented model selection using Akaike’s Information Criterion (AIC (16)). AIC is a standard for comparing the goodness-of-fit of models whilst accounting for their respective complexity, and can be used for all models for which a likelihood can readily be calculated. It is defined as:

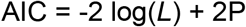

where *L* is the model likelihood (so that –2 log*(L)* is the model’s deviance) and *P* is the number of parameters of the model. AIC is calculated for each model, and the model with lowest AIC is retained as the ‘best fitting’ model. This procedure is fast but it does not guarantee that the retained model has the best predictive abilities. In practice though, results on simulated data suggest both approaches often give identical results (results not shown), in line with recent observations on incidence forecasting (24).

#### Setting the prediction window (k)

The length of the prediction window *k* is an important parameter in ASMODEE, as it draws the line between the time period used for estimating past trends, and the recent time period over which we identify potential anomalies. For routine surveillance, a possible approach is to fix arbitrary values for both the calibration window and the prediction window, e.g. using 7 weeks of daily incidence data for calibration and looking for outliers in the last week of data. These values may need to be adjusted over time to ensure optimal detection of changes in temporal trends, and to balance the need for the calibration window to contain sufficient data points to fit the most complex time series model considered. In addition, the value of *k* should be chosen so that i) it exceeds delays previously identified for detecting trend changes and ii) it remains within the time period over which forecasts are deemed reliable (typically no more than 3 weeks for fast-spreading diseases such as Ebola or COVID-19).

Besides this ‘manual tuning’ approach, heuristics can be used to compare results over different values of *k*. Various criteria can be devised to optimise *sensitivity* (ability to detect trend changes) or *specificity* (ability to identify data points following the trend), or any trade-off of these. Here, we maximise a simple score calculated for each value of *k* (up to a user-specified maximum value) as the sum of two components: i) the number of non-outliers (points within the prediction interval of the ‘best’ model) in the calibration window, and ii) the number of outliers in the prediction window. Therefore, this criterion tends to select a value of *k* such that past temporal trends can be well-defined (i.e. with most pre-break points within the prediction interval), whilst also retaining the ability to detect recent anomalies.

### Application to simulated data

#### Simulated data

ASMODEE was evaluated using simulated COVID-19 incidence. All simulations used the *projections* package version 0.5.1 (25), which implements branching-process epidemic simulations allowing for the reproduction number *(Rt)* to vary by time periods. A full description of the model can be found in Jombart *et al*. (26). Note that here and below, *Rt* refers to the expected number of secondary cases, and not the actual number of secondary infections drawn, so that even with *Rt* = 1 some simulated epidemic trajectories might increase or decrease over time. Briefly, COVID-19 infections are simulated on a given day from a Poisson distribution, according to a force of infection determined by past cases, and distributions of the serial interval and *Rt*. Here, we used a Gamma-distributed serial interval with mean 4.7 days and standard deviation 2.9 days (27). *Rt* was assumed to be normally distributed with different means for different scenarios, and standard deviations 10% of the mean. We accounted for under-reporting by assuming that on average 10% of infections were reported (this value is broadly compatible with reported estimates (28)), so that the observed case count followed a binomial distribution with the number of trials the true number of cases, and probability of success of 0.1.

We considered four scenarios which translated into different initial incidences and time-varying values of *Rt*:

- *Steady state:* 1,000 cases per day (thus an average of 100 reported cases per day) with a mean *Rt* of 1.
- *Relapse:* 10,000 initial cases per day first declining with mean *Rt* of 0.8, followed by an increase with a mean *Rt* of 1.3.
- *Lockdown:* 1,000 initial cases per day first increasing with a mean *Rt* of 1.3, followed by a decline with mean *Rt* of 0.8.
- *Flare-up:* 10 initial cases per day and a steady state with a mean *Rt* of 1, followed by a sudden increase with mean *Rt* 2.5.

All simulations produced daily time series for 42 days. An *observation period*, over which the algorithms were evaluated, was set to be the 12 last days. This period, a bit longer than a typical use case of 7 days, was chosen to offer enough variability and data points for a meaningful evaluation. Changes in *Rt* in the last three scenarios took place on days 34, 35 or 36. Simulations were replicated 30 times for each scenario and day of period change, resulting in 30 simulated time series for the *steady state* scenario and 90 for the others (Figure 1).

**Figure 1.**
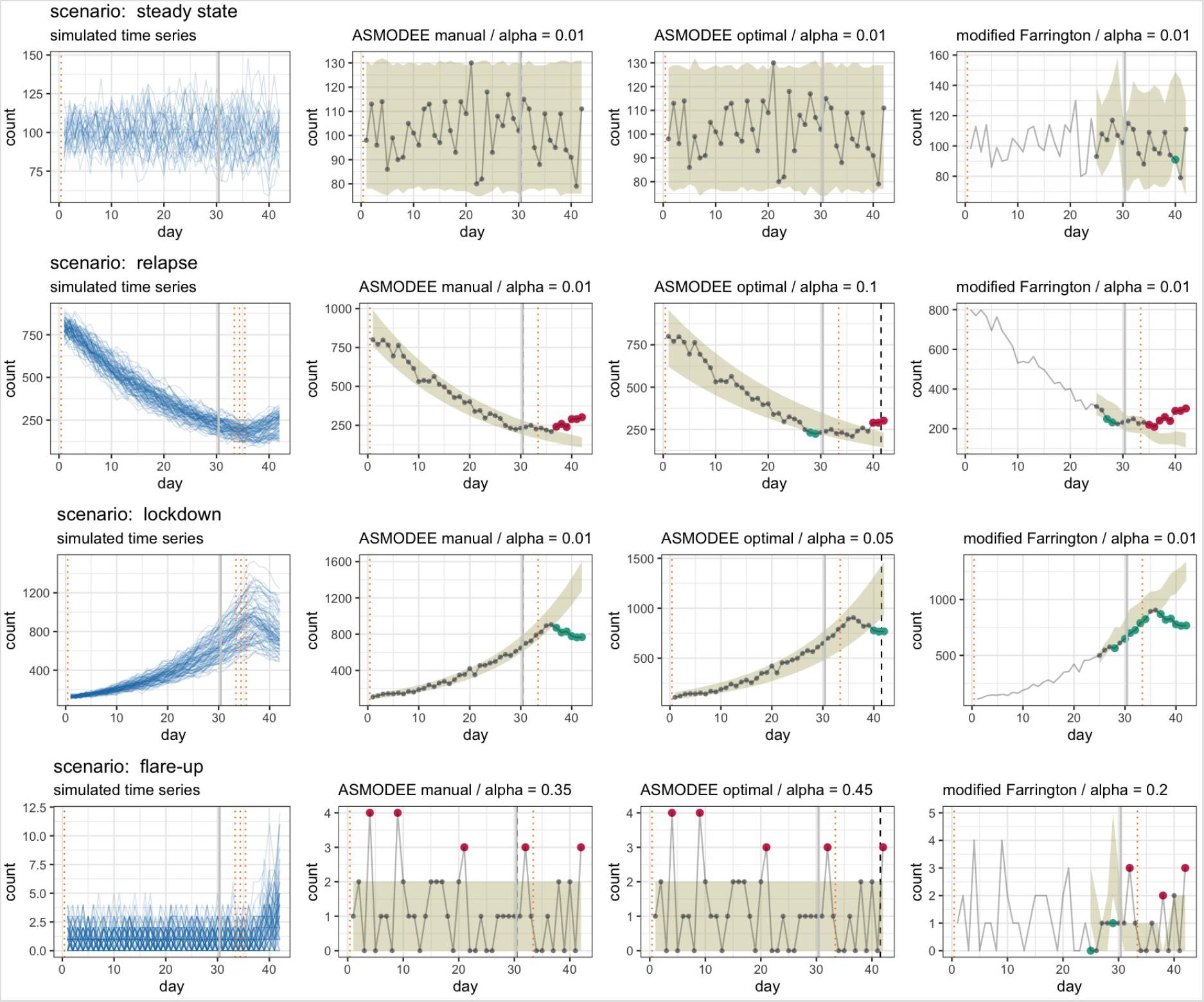
Simulations under four scenarios and illustration of the detection algorithm. Left column: The blue lines show time series generated under the four scenarios *steady state* (30 simulations), *relapse, lockdown* and *flare-up* (90 simulations each). The solid grey line marks the beginning of the observation period, the dotted orange lines the time at which a trend change occurs. Second to fourth columns: application on a single time series from each scenario of the configurations *ASMODEE manual, ASMODEE optimal*, and the *modified Farrington* algorithms. The grey lines show the simulated time series, the ribbon shows the prediction interval for the value of *alpha* indicated, and the dashed black line marks the end of the calibration period for ASMODEE. The red dots show data points categorised as *increase*, the green dots those categorised as *decrease*, the grey ones are considered *normal*. The values of *alpha* were set so as to maximise the six-day probability of detection. *modified Farrington* does not produce results for the first training period.

#### Outbreak detection

Here, we propose a general framework to evaluate COVID-19 outbreak detection algorithms. We used it to compare two configurations for ASMODEE together with the *modified Farrington* algorithm as a benchmark.

ASMODEE was applied to the simulated time series with one of three models (Poisson GLM with constant mean, linear regression with temporal effect, negative binomial GLM with log-linear trend in time) selected through minimization of the AIC. Model selection through cross-validation produced identical results (data not shown). We looked at either a fixed *k* of 12 days *(ASMODEE manual)* or an optimal *k* of at most 12 days *(ASMODEE optimal)*. The rationale for setting *k* to (at most) the duration of the observation period is that a user would define a period in which they are interested, implicitly assuming everything that happened before was not of direct interest and thus could be used to train the algorithms.

The *modified Farrington* algorithm (14) trains a statistical model on univariate count time series to derive bounds based on a parameter *alpha*. It takes trend and seasonality into account and downweights anomalous counts in the training range. To avoid getting caught in recent trends and thus miss outbreaks, the last time steps are ignored during training. Conversely, low counts can be ignored to avoid uninteresting alarms. Lastly, it performs a one-step ahead detection, i.e. it is retrained at every time step. It is typically used for weekly detection of point-source outbreaks with comparison to a five-years baseline, but its parameters can be adapted to reflect other situations. Here, we use its implementation as farringtonFlexible in the R package *surveillance* (9) with three weeks training of a GLM with Negative Binomial family, weekly periodicity and a three-days window half-size around the first day of each period, no limitation on low counts, and the last week ignored for training. Moreover, we further adapted the algorithm so that it always takes trend into account, thus removing the requirement that the estimated mean not be higher than all preceding counts.

The algorithms classified each day of a given time series as either *increase, decrease* or *normal* if it lied above the upper bound of the prediction interval, below its lower bound, or between the two, respectively. The application of the algorithms and the resulting classifications are illustrated for each scenario in Figure 1.

#### Evaluation

To evaluate the three approaches (*ASMODEE manual, ASMODEE optimal* and *modified Farrington)*, we needed to define the expected ‘truth’ to which the classifications were compared and to derive scores relevant for the practical use. First, we translated trend periods into properties of each day: all points before a trend change, or if there was no change, were considered *normal*; if the trend changed upwards, i.e. if the mean *Rt* of the second period was larger than in the first, all subsequent days were considered an *increase*; conversely, a decline in mean *Rt* led to subsequent days being considered *decrease*. Thus, the detection task could be framed as a multiclass classification. To reflect practical use, only the days within the observation period were taken into account for the evaluation.

In the four scenarios considered this simplified to binary classifications, as at most two classes were present, with the last period defining the positive class: days could thus be correctly classified as ‘positive’ *(i.e*. change of trend; we denote the number of these true positives by TP), wrongly as positive (FP, the number of false positives), correctly as ‘negative’ *(i.e*. no change of trend, TN) or wrongly as negative (FN). From those, standard scores can be computed for each time series, of which we considered the following:

1. *sensitivity* TP/(TP+FN);
2. *specificity* TN/(TN+FP);
3. *balanced accuracy, ba* = (sensitivity+specificity)/2;
4. *precision* TP/(TP+FP); and *F1 score*,
5. *F1* = 2⋅precision⋅sensitivity/(precision+sensitivity).

The value these scores take depends on *alpha*, which was set to an optimal value as described below.

In contrast to these daily classifications, we also defined the *period detection*, which indicated whether a true change was detected over a given time period. This led to the definition of a *probability of detection within a time interval D* as the proportion of time series with the last period correctly detected within *D* days after the trend change. It thus represents a measure of *timeliness* of trend-change detection. Figure 3 shows the probability of detection computed for each scenario, algorithm and selected values of the threshold parameter *alpha*.

*alpha* can be seen as an adjustment parameter: the smaller its value, the higher the proportion of days/periods classified as *normal*. For the purpose of comparing and evaluating algorithms we varied *alpha* from 0.01 to 0.5. Its optimal value was defined as the one maximizing the probability of detection within 6 days, the longest interval available for all simulation runs (Figure 3). In practice, *alpha* can be iteratively adjusted by the user to achieve the desired level of alert for a given surveillance system.

### Automated pipelines for NHS Pathways data

#### Implementation

As a real-world use case we used the NHS Pathways data (29) reporting potential COVID-19 cases in England, broken down by Clinical Commissioning Groups (CCG). These data include all reports classified as ‘potential COVID-19 cases’ notified via calls to 111, 999, and 111-online systems (30). These data are not confirmed cases, and are subject to unknown reporting biases. They likely include a substantial fraction of ‘false positives’ (cases classified as potential COVID-19 which are in fact due to other illness), as well as under-reporting (true COVID-19 cases not reported). Last, as these data are using self-reporting, it is likely that individual perceptions as well as ease of access to the reporting platforms impact the observed numbers.

We have discussed how these data can be interpreted and the associated caveats elsewhere and refer to these sources for more context (31). Recent observations suggest that 111/999 calls may be more reflective of COVID-19 dynamics than 111-online (31). Therefore, the subsequent analyses were based on these data only.

We have developed an automated pipeline to download these data, apply ASMODEE and present results. It has been implemented in a publicly accessible webpage (see below the section ‘Availability’) and was used in the illustration discussed in the next section.

#### Illustration

The pipeline for NHS Pathways data was applied to all CCG’s each day from 1st June to 10 August 2020. We highlighted the results for Leicester City and Blackburn as they experienced a large COVID-19 outbreak in the middle of June and July, respectively, with the first leading to the first local lockdown in the UK being imposed on 29 June.

We chose as parameters for ASMODEE a fixed *k* of 7 days and used AIC as the criterion for model selection. Here also, model selection through cross-validation produced identical results (data not shown). *alpha* was set to 0.05, low enough to avoid false alarms but high enough to rapidly detect changes. The five candidate models from which one was selected each day were: Poisson with constant mean; linear regression with linear trend in time; negative binomial with log-linear trend in time; the same with supplementary co-factor the day of the week (distinguishing weekends, Mondays, and other days, in order to account for system closure over weekends and back-log effects); and the latter with a supplementary interaction of day and day of the week. Figure 4 shows the output of ASMODEE as applied each day, using a calibration window of 35 days. Moreover we considered one metric to compare CCG’s and highlight the peculiarity of the situations in Leicester and Blackburn with Darwen: the number of *increase* signals in a 7-day observation period (Figure 5).

### Availability

#### ASMODEE

ASMODEE is implemented in the new package *trendbreaker* for the R software (32), released under MIT license as part of the toolkit developed by the R Epidemics Consortium (RECON, http://repidemicsconsortium.org/) for outbreak analytics (33). Whilst already functional and fully documented, this package is still under active development, and will be an integral part of the next generation of tools for handling, visualising and modelling epidemic curves developed within RECON. It is available and documented at: https://github.com/reconhub/trendbreaker, and is scheduled for a release on CRAN by the end of 2020.

The package provides a unified interface for various modelling tools including linear models (function lm), Gaussian, Binomial, quasi-Binomial, Poisson, quasi-Poisson and Gamma GLMs (function glm), negative binomial GLM (function MASS::glm.nb) and Bayesian generalised non-linear models using Stan (34,35) (function brms::brm). Various routines are implemented for automated model selection (function select_model), outlier detection (function detect_outliers) and the selection of *k* (function detect_changepoint). The main function asmodee wraps these different tools and provides a full implementation of the method, including the two approaches for model selection (repeated *K*-fold cross-validation and AIC) and the automated selection of *k* using the scoring described in the previous section, plus plotting functions.

#### Simulations

Scripts implementing the simulation of COVID-19 outbreaks, their analysis as well as the adapted farringtonFlexible function are available at: https://gitlab.com/stephaneghozzi/asmodee-trendbreaker-evaluation.

#### NHS Pathways analysis pipeline

The data pipeline applying ASMODEE to NHS Pathways data is available at: https://github.com/thibautjombart/nhs_pathways_monitoring. This pipeline is designed as a blogdown website (36) which automatically updates data and analyses daily using github actions. Daily data cleaning itself is done by the data pipeline hosted at: https://github.com/qleclerc/nhs_pathways_report. The resulting website renders through Netlify at: https://covid19-nhs-pathways-asmodee.netlify.app/. Therefore, our pipeline is entirely free and can easily be replicated and adapted to other data streams. The script for illustration on CCG data are available at: https://gitlab.com/stephaneghozzi/asmodeetrendbreaker-evaluation.

## RESULTS

### Simulated data

ASMODEE detected anomalies in the three scenarios with trend changes in the simulated time series for both the manual and optimal tuning (Figure 1). Very few false alarms were generated in the *steady state* scenario.

The five performance scores varied substantially within and across scenarios. All approaches showed almost perfect sensitivity in the *steady state* scenario, where typically more than 99% of ‘normal’ days are classified as such. Results were more heterogeneous in the *relapse* and *lockdown* scenarios where both ASMODEE configurations showed high precision and specificity, with varying sensitivity, translating into high overall balanced accuracy and F1 scores (Figure 2). *ASMODEE manual* performed somewhat better in sensitivity and thus in F1 scores. Both ASMODEE approaches performed similarly in the *flare-up* scenario. Overall, the methods exhibited high specificity and precision, but with lower sensitivity than in the *lockdown* and *relapse* scenarios. *modified Farrington* performed very similarly to *ASMODEE manual*, with slightly higher sensitivity and lower specificity in the *lockdown* scenario.

**Figure 2.**
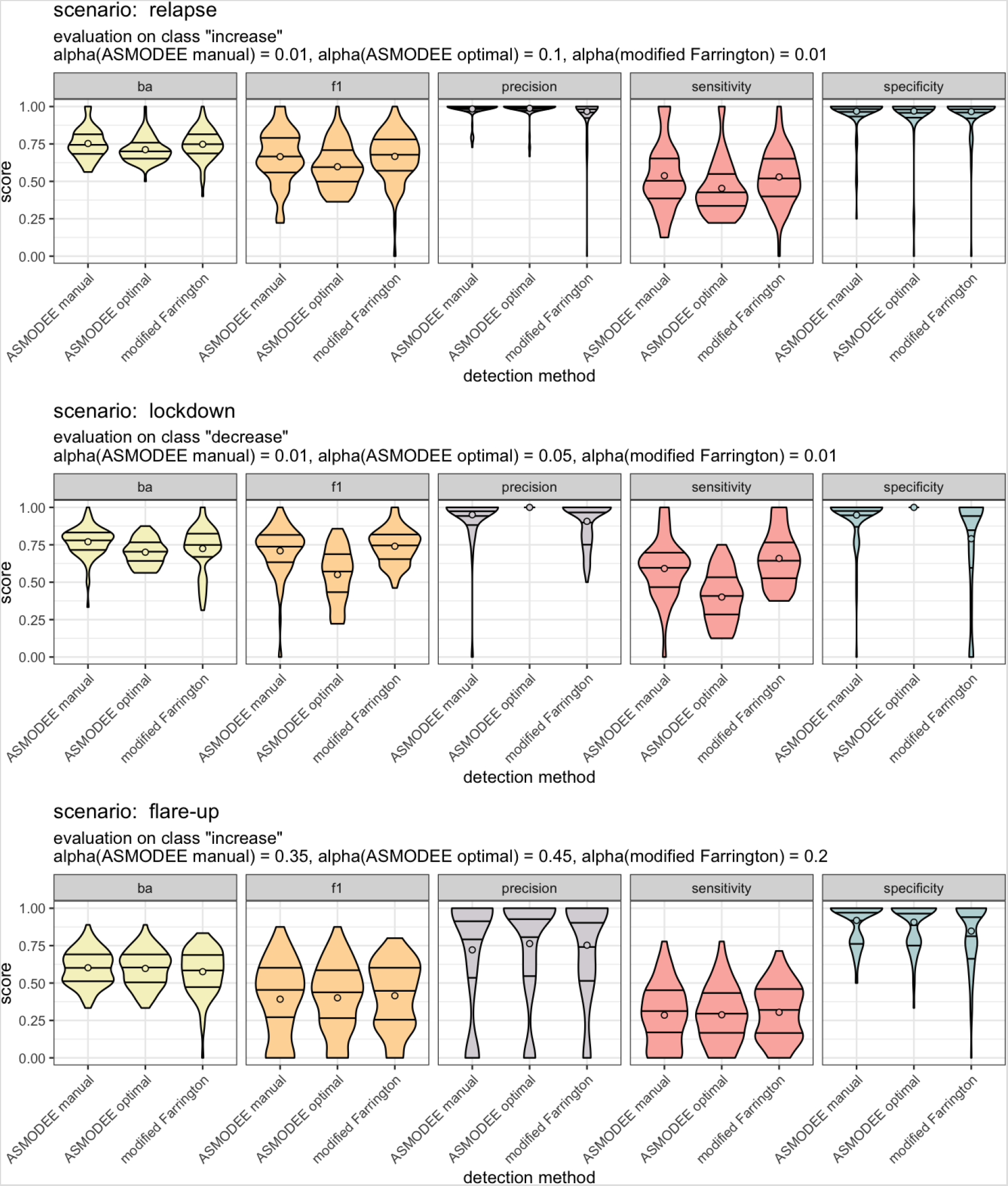
Evaluation of the algorithm on simulated data. Distribution of scores over all simulation runs. Each row corresponds to a different scenario, each column to one score, computed for each algorithm. “ba” stands for balanced accuracy, the average of sensitivity and specificity. The horizontal lines correspond to the 25th, 50th and 75th percentiles, the dots correspond to the average. The plot title indicates which class was considered positive for each scenario and which value of *alpha* was considered optimal. (The trivial results for the *steady state* scenario, with a sensitivity very close to 1 and other scores undefined, are not shown.)

It should be noted that results are not only a reflection of the methods evaluated, but are also largely conditioned by the simulations and evaluation settings. Because we used a branching process to simulate COVID-19 outbreaks, changes in *Rt* are typically reflected after at least one serial interval (4 to 5 days on average). The resulting lag in the incidence time series, which can be seen in Figure 1, imposes an upper bound on sensitivity: algorithms can only detect changes in transmissibility once these are reflected in the case counts. With the last trend period encompassing 7 to 9 days in our simulations, around half of the data points at best can be expected to be correctly detected as anomalies.

The probability of detection within 6 days or less is less sensitive to the choice of observation period, as it does not depend on its duration. In Figure 3 shows, *alpha* has a non-monotonic effect on the detection performance, leading to an optimal value maximised after 6 days. In the three scenarios with a trend change a 50% probability of detection for ASMODEE is typically reached only after 3 to 5 days, again due to the delay between changes in *Rt* and its actual reflection in the incidence time series. The detection was less successful in the *flare-up* scenario as very low, on average constant numbers and sudden increase exacerbated the effect of delays. Overall, in contrast to sensitivity, *ASMODEE optimal* tended to achieve a better probability of detection than *ASMODEE manual*, reaching almost 100% in the *relapse* and *lockdown* scenarios. In contrast, *modified Farrington* showed lower probabilities of change detection across the values of alpha considered for the *increase* and *lockdown* scenarios.

**Figure 3.**
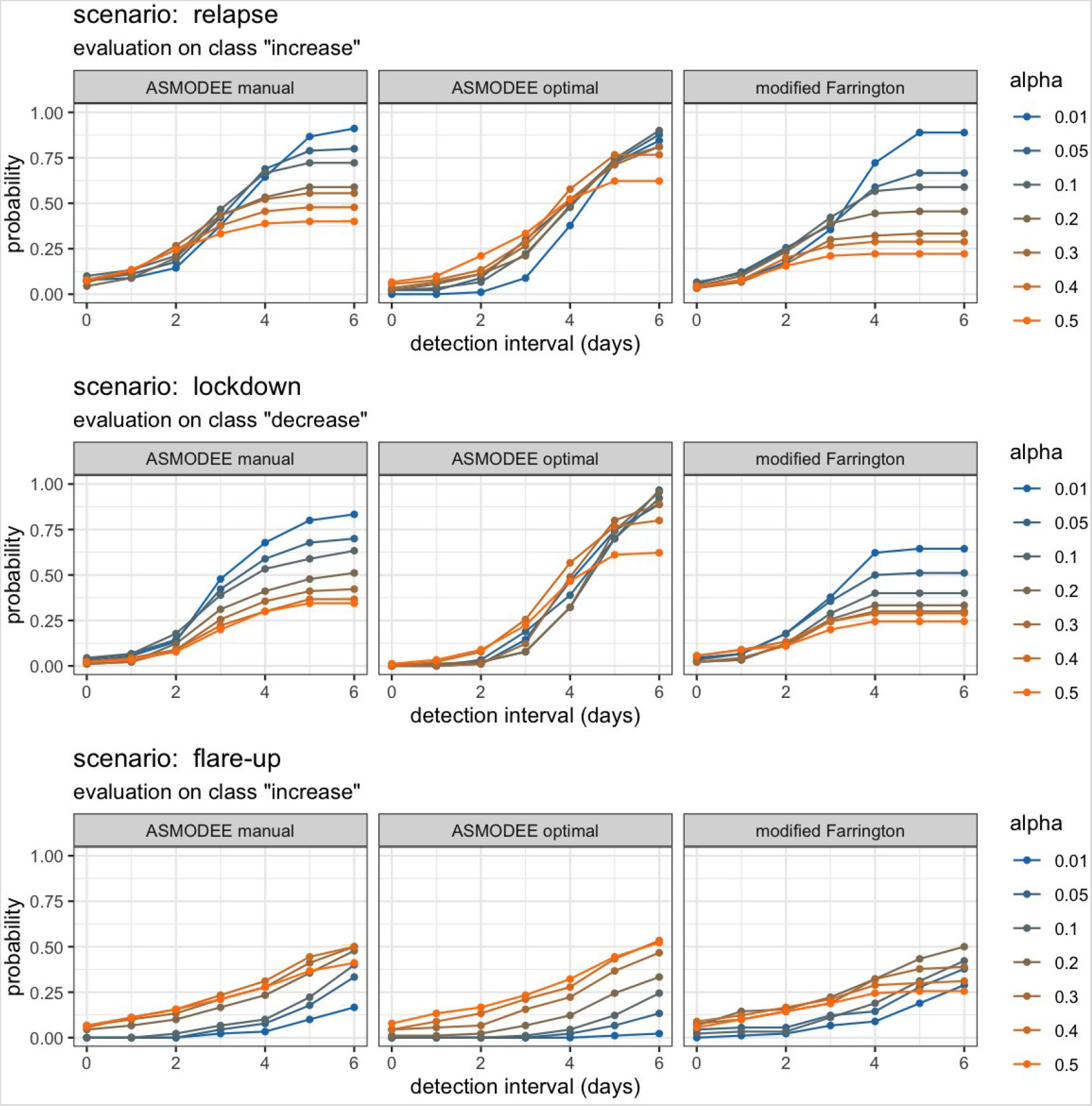
Delay to trend change detection. This figure illustrates the delay between the change in *Rt* and the first detected change *(increase* or *decrease)* by the different methods. The probability of detection on the y-axis is defined as the proportion of detection for a given delay, across all simulations. Results are provided for selected values of the threshold parameter *alpha*, represented in color. (The trivial results for the *steady state* scenario are not shown.)

### Detection of flare-ups of COVID-19 cases in England

Looking at the particular case of Leicester in June 2020, we see that *increase* signals are consistently triggered for reference dates ranging from 10 to 22 June (Figure 4). ASMODEE indicates 1 anomalous day from 7–9 June, two on 10 June, and three on 11 June. Consistent with simulation results, ASMODEE exhibited good specificity, with very few *increase* signals generated before or after the relapse. Similar patterns were observed for Blackburn from 11 until 18 July 2020 (data not shown). The algorithm correctly flagged the weeks following the lockdown in Leicester on 29 June and the associated drop in incidence as *decreases* (Figure 4).

**Figure 4.**
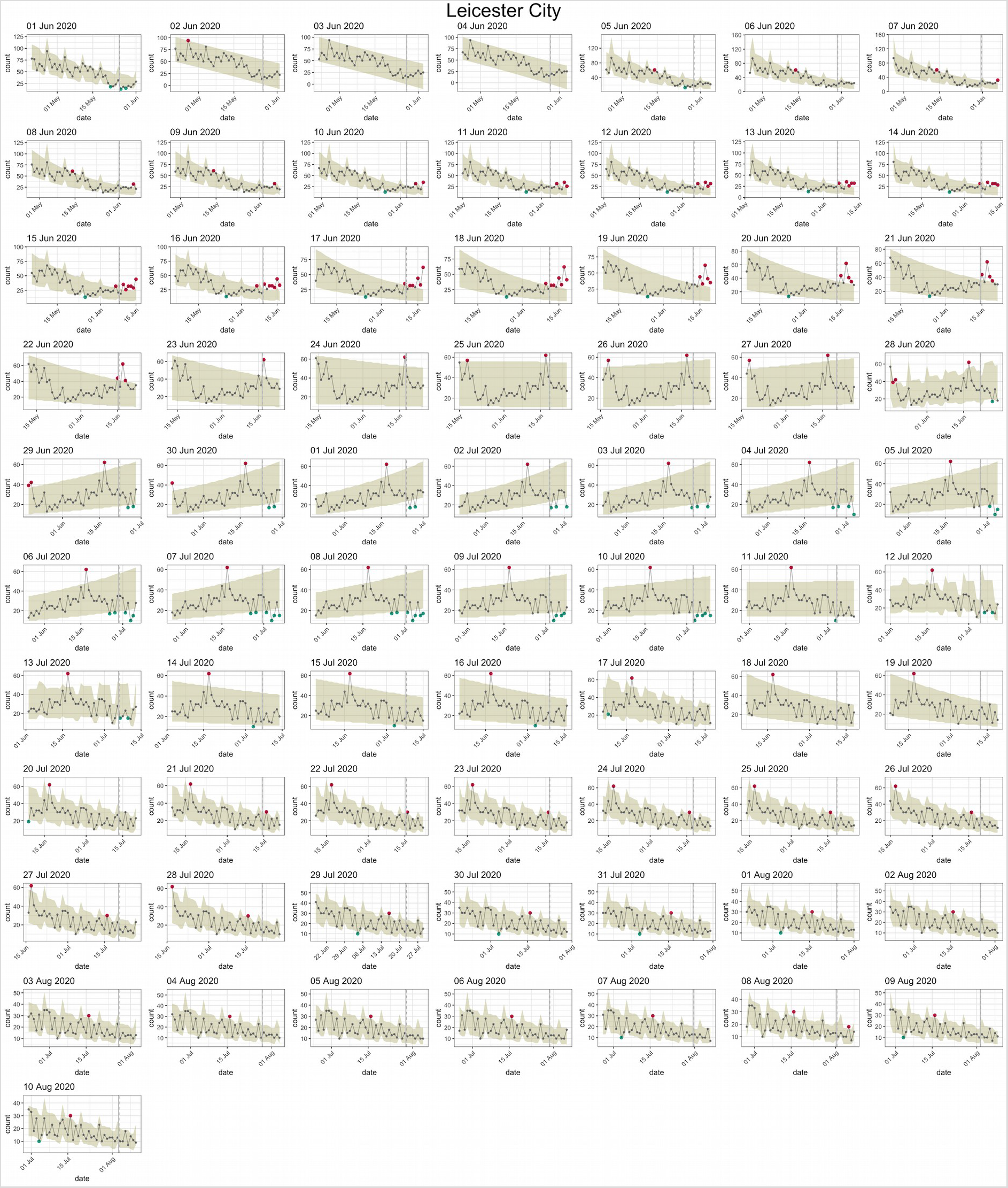
ASMODEE applied to the COVID-19 cases reported through the NHS Pathways (111/999 calls) for the CCG Leicester City during June and July 2020. Each plot corresponds to a different reference date, with the algorithm applied to the preceding 42 days (including the reference day). *k* was fixed to 7 days, *alpha* was set to 0.05. Considered temporal trends models included: Poisson GLM with constant mean, linear regression with linear trend in time, negative binomial GLM with log-linear trend in time, the same with a ‘day of the week’ effect (distinguishing weekends, Mondays, and other days), and the latter with an additional interaction to allow for different slopes by day of the week. Automated model selection using AIC was used for all analyses.

To understand the context of the increase in the Leicester and Blackburn with Darwen CCG’s, we analysed the time series of cases in all 136 CCGs which reported potential COVID-19 cases in June and July 2020. Leicester showed one of the longest periods of consecutive increase from 13 to 21 June (Figure 5). Similarly, Blackburn stands out from 13 to 18 July. Taken together, these results show that ASMODEE could have detected the outbreaks in both regions early on.

**Figure 5.**
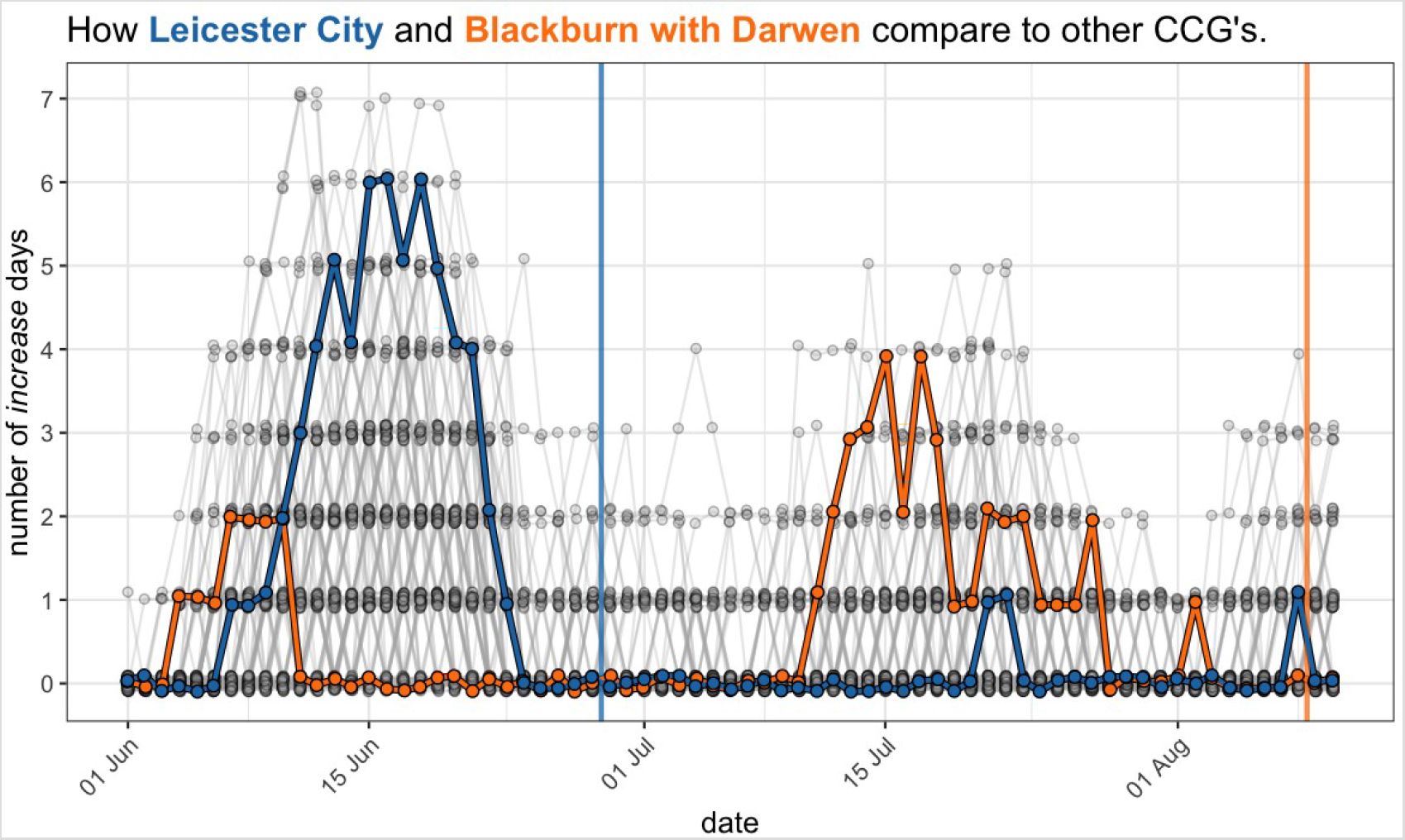
Leicester City and Blackburn with Darwen stand out among all CCG’s from 1st June until 10 August 2020. Number of days classified as *increase* within 7 days of the reference date. Each dot represents one CCG on one reference day, the lines connect the dots corresponding to the same CCG’s from one day to the next. Leicester City is highlighted in blue, Blackburn with Darwen in orange. A small jitter was applied for better readability. The blue and orange vertical lines indicate the dates at which increased restrictions were imposed in Leicester and Blackburn, respectively.

## DISCUSSION

We have presented ASMODEE, a novel algorithm for trend-change and anomaly detection. ASMODEE unifies two important features of an early warning system: gauging temporal trends in incidence time series, and detecting recent anomalies, exceptionally higher case counts than expected based on recent data. The main advantage of the approach may be its flexibility for modelling temporal trends. Rather than imposing a single model, it allows for a range of candidate models to be specified, and automatically selects the best predicting (using repeated *K*-fold cross validation) or fitting (using AIC) model. The current implementation provides support for linear regression, various different types of GLMs GLMs, as well as Bayesian regression implemented in Stan, and should as such provide ample flexibility for modeling different epidemic time series.

We tested ASMODEE using simulated COVID-19 epidemics, but this approach can be readily extended to other diseases and contexts. For instance, ASMODEE could be useful for detecting new hotspots of Ebola cases in large-scale epidemics such as the West-African Ebola outbreak (37,38) or the more recent one in North Kivu / Ituri, Democratic Republic of the Congo (26,39). In a different context, it could complement existing systems for tracking nosocomial outbreaks of norovirus at a national level (40), to help identify wards or hospitals with a sudden increase of cases which would warrant prompt intervention.

We compared two variants of ASMODEE (with or without automatically setting the parameter *k)* to a state-of-the-art outbreak detection algorithm in *modified Farrington*. Considering a number of metrics that reflect how a user would typically interpret detection results, we found that all three methods performed overall well, with very high specificity and varying sensitivity. Using a fixed value of *k* in ASMODEE (‘*ASMODEE manual*’) seemed to increase sensitivity in the considered scenarios. As this approach is also more computationally efficient, it may be a reasonable default for setting up surveillance pipelines. The *ASMODEE manual* configuration performed very closely to *modified Farrington*. This similarity may not be surprising: without seasonality and past, localised outbreaks, both approaches are based on similar statistical modeling and aberration detection approaches. Interestingly, when considering the delay to change detection, ASMODEE optimal showed more consistent, and better performances across different values of alpha than the other two methods in two scenarios out of three. Importantly, the low sensitivity exhibited in some results can be largely explained by the branching process used to simulate outbreaks: changes in *Rt* are only actually reflected in incidence time series after a lag of at least one serial interval. Further work may focus on evaluating ASMODEE using alternative models to simulate time series without such lag.

ASMODEE, as implemented in the package *trendbreaker*, is both much simpler than *modified Farrington* (requiring fewer parameters to be fine tuned for specific contexts and diseases) and much more flexible (many kinds of statistical models can be easily integrated and automatically selected), as illustrated by the fact that we had to adapt *modified Farrington* to account for upward trends and produce meaningful results. However, it is not the case that ASMODEE would always be the approach of choice: *modified Farrington* has been developed for very specific use cases and practical needs, namely the longer term surveillance of infections causing point-source outbreaks, such as food- or water-borne diseases. Nevertheless, we believe it to be a suitable benchmark for COVID-19 as, for lack of more specific standard tools, it would still have been the default method employed for COVID-19 surveillance.

Other aspects of the ASMODEE remain to be investigated. In our simulations, model selection using repeated *K*-fold cross validation and AIC gave near identical results (results not shown). This may not be the case in other settings, and further work will be needed to investigate the automated model selection step. In particular, one may look for optimal fold sizes and numbers of repeats in the cross validation procedure, and may also consider other goodness-of-fit statistics such as the Bayesian Information Criteria (BIC(17)). Further developments may also be considered for defining the optimal value *k*, beyond the simple scoring system introduced here.

The proposed method has a number of limitations. The main one relates to reporting delays, which are an intrinsic feature of most epidemiological data. ASMODEE takes incidence data on face value, *i.e*. without accounting for the potential effect of reporting delays, which typically cause incidence time series to artificially decrease over the last few days of data. While this limitation is not specific to ASMODEE, it will clearly hinder the method’s capacity to detect recent increases in case counts. A possible improvement of the method would be to characterise reporting delays, and then use augmented data / nowcasting to simulate the true underlying incidence, on which ASMODEE would be run. This approach would undoubtedly increase computational time, but would be easy to parallelise and most likely still fast enough to be used in daily surveillance of hundreds of geographic locations.

A second limitation of our approach is that ASMODEE does not consider spatial spread of epidemics. While multiple locations can be analysed separately as illustrated in our analysis of NHS 111/999 data, the approach does not account for transmission across different locations. The general framework used in ASMODEE could in theory be extended to multivariate time series models incorporating spatial dependency, but the current implementation would need additional work to support such features. In practice, we expect this may only be a substantial limitation when very good data on patient locations and movement is available. An alternative worth exploring would be to run ASMODEE on spatially smoothed data (41) to reduce noise in data typically observed in small geographic areas with low case numbers and therefore facilitate anomaly detection.

We expect ASMODEE will be most useful for surveying potentially large numbers of incidence time series, *e.g*. to detect flare-ups of COVID-19 in small geographic units and/or specific population demographics. It may be best used in conjunction with human judgment rather than as a purely automated algorithm. For instance, ASMODEE could rank incidence times series according to their respective numbers of ‘increase’ days, and then the highest ranked series could be examined by epidemiologists to decide whether further investigation is warranted. As such, ASMODEE could form the basis for a daily COVID-19 surveillance system, and be regularly refined *e.g*. changing the duration of the detection window (parameter *k)* or the alpha threshold to meet required alert levels. The automated pipeline we have developed for NHS 111/999 data shows that such surveillance systems can be built using free, open-source tools, and readily automated for daily updates.

It is our hope that ASMODEE will form a useful complement to existing point-source outbreak detection methods such as the *modified Farrington* algorithm (14) and scan statistics (42). We believe its inherent simplicity and flexibility for modelling time series, together with its availability in the free, open-source R package *trendbreaker*, will facilitate further improvements and adoption by the community, and its broader use for disease surveillance for COVID-19 outbreaks and beyond.

## Data Availability

All scripts and data are publicly available on github (see links in the manuscript).

https://github.com/reconhub/trendbreaker

https://gitlab.com/stephaneghozzi/asmodee-trendbreaker-evaluation

https://github.com/thibautjombart/nhs_pathways_monitoring

https://covid19-nhs-pathways-asmodee.netlify.app/

## ACKNOWLEDGEMENTS

The named authors (OB, JE, RE, TJ, MJ, SM, EN) had the following sources of funding: OB was funded by a Sir Henry Wellcome Fellowship funded by the Wellcome Trust (206471/ Z/17/Z). RE receives funding from HDR UK (grant number: MR/S003975/1). SF is supported by a Sir Henry Dale Fellowship jointly funded by the Wellcome Trust and the Royal Society (Grant number 208812/Z/17/Z). TJ receives funding from the Global Challenges Research Fund (GCRF) project ‘RECAP’ managed through RCUK and ESRC (ES/P010873/1), the UK Public Health Rapid Support Team funded by the United Kingdom Department of Health and Social Care and from the National Institute for Health Research (NIHR) – Health Protection Research Unit for Modelling Methodology. MJ receives funding from the Bill and Melinda Gates foundation (grant number: INV-003174) and the NIHR (grant numbers: 16/137/109 and HPRU-2012–10096). EN receives funding from the Bill and Melinda Gates Foundation (grant number: OPP1183986). MJ and JE receive funding from European Commission project EpiPose (101003688). SM receives funding from the Wellcome Trust (grant: 210758/ Z/18/Z). SFlasche receives funding from the Wellcome Trust (grant: 208812/Z/17/Z).

The following funding sources are acknowledged as providing funding for the working group authors. Alan Turing Institute (AE). BBSRC LIDP (BB/M009513/1: DS). This research was partly funded by the Bill & Melinda Gates Foundation (INV-001754: MQ; INV-003174: KP, MJ, YL; NTD Modelling Consortium OPP1184344: CABP, GFM; OPP1180644: SRP; OPP1183986: ESN; OPP1191821: KO’R, MA). BMGF (OPP1157270: KA). DFID/Wellcome Trust (Epidemic Preparedness Coronavirus research programme 221303/Z/20/Z:KvZ). DTRA (HDTRA1–18–1–0051: JWR). Elrha R2HC/UK DFID/Wellcome Trust/This research was partly funded by the National Institute for Health Research (NIHR) using UK aid from the UK Government to support global health research. The views expressed in this publication are those of the author(s) and not necessarily those of the NIHR or the UK Department of Health and Social Care (KvZ). ERC Starting Grant (#757699: JCE, MQ, RMGJH). This project has received funding from the European Union’s Horizon 2020 research and innovation programme - project EpiPose (101003688: KP, MJ, PK, RCB, YL). This research was partly funded by the Global Challenges Research Fund (GCRF) project ‘RECAP’ managed through RCUK and ESRC (ES/P010873/1: AG, CIJ, TJ). HDR UK (MR/S003975/1: RME). NIHR (16/136/46: BJQ; 16/137/109: BJQ, CD, FYS, MJ, YL; Health Protection Research Unit for Immunisation NIHR200929: NGD, FGS; Health Protection Research Unit for Modelling Methodology HPRU-2012–10096: TJ, FGS; NIHR200929: MJ; PR-OD-1017–20002: AR). Royal Society (Dorothy Hodgkin Fellowship: RL; RP\EA\180004: PK). UK DHSC/UK Aid/NIHR (ITCRZ 03010: HPG). UK MRC (LID DTP MR/N013638/1: GRGL, QJL, NRW; MC_PC_19065: AG, NGD, RME, SC, TJ, YL; MR/P014658/1: GMK). Authors of this research receive funding from UK Public Health Rapid Support Team funded by the United Kingdom Department of Health and Social Care (TJ). Wellcome Trust (206250/Z/17/Z: AJK, TWR; 206471/Z/17/Z: OJB; 208812/Z/17/Z: SC, SFlasche; 210758/Z/18/Z: SFunk, JM, SA, JH, NIB). No funding (AKD, AMF, CJVA, DCT, SH, YWDC).

The UK Public Health Rapid Support Team is funded by UK aid from the Department of Health and Social Care and is jointly run by Public Health England and the London School of Hygiene & Tropical Medicine. The University of Oxford and King’s College London are academic partners. The views expressed in this publication are those of the authors and not necessarily those of the National Health Service, the National Institute for Health Research or the Department of Health and Social Care.

## AUTHORS CONTRIBUTIONS

In alphabetic order:

SG, TJ, DS developed the methodology.

SG, TJ, DS, contributed code.

SG, TJ, performed the analyses.

SG reviewed code.

SG, TJ, wrote the first draft of the manuscript.

OB, JE, RE, SF, FG, SG, MH, TJ, MJ, QL, GM, SM, EN, DS, TW contributed to the manuscript.

CMMID COVID-19 Working Group gave input on the method, contributed data and provided elements of discussion. The following authors were part of the Centre for Mathematical Modelling of Infectious Disease 2019-nCoV working group: Arminder K Deol, Kathleen O’Reilly, Charlie Diamond, David Simons, Petra Klepac, Christopher I Jarvis, Sebastian Funk, Nicholas G. Davies, Yung-Wai Desmond Chan, Damien C Tully, Nikos I Bosse, Simon R Procter, Kaja Abbas, Amy Gimma, Jon C Emery, Billy J Quilty, Kevin van Zandvoort, Stéphane Hué, Rosanna C Barnard, Timothy W Russell, Sam Abbott, Kiesha Prem, Adam J Kucharski, Akira Endo, Fiona Yueqian Sun, James W Rudge, Katharine Sherratt, Yang Liu, Katherine E. Atkins, Rein M G J Houben, Matthew Quaife, Joel Hellewell, Gwenan M Knight, Carl A B Pearson, Georgia R Gore-Langton, Anna M Foss, Megan Auzenbergs, Alicia Rosello, Samuel Clifford, C Julian Villabona-Arenas, Hamish P Gibbs, Alicia Showering, Jack Williams, Frank G Sandman, Naomi R Waterlow. Each contributed in processing, cleaning and interpretation of data, interpreted findings, contributed to the manuscript, and approved the work for publication.

## Notes

### Competing Interest Statement

The authors have declared no competing interest.

### Author Declarations

N/A: this work only uses publicly available data

